# Long-term durability and public health impact of city-wide *w*Mel *Wolbachia* mosquito releases in Niterói, Brazil during a dengue epidemic surge

**DOI:** 10.1101/2025.04.06.25325319

**Authors:** Katherine L Anders, Gabriel Sylvestre Ribeiro, Renato da Silva Lopes, Pilar Amadeu, Thiago Rodrigues da Costa, Thais Irene Souza Riback, Karlos Diogo de Melo Chalegre, Wesley Pimentel de Oliveira, Cátia Cabral da Silva, Marcos B Vinicius Ferreira Mendes Blanco, Ana Lucia Fontes Eppinghaus, Fabio Villas Boas, Tibor Frossard, Benjamin R Green, Scott L O’Neill, Peter A Ryan, Cameron P Simmons, Luciano Andrade Moreira

## Abstract

In 2024, the Americas region experienced the largest dengue outbreak on record and Brazil was among the worst affected countries, reporting 6.6 million cases and 6,200 deaths. We report the long-term entomological and epidemiological effectiveness of city-wide deployment of *w*Mel-strain *Wolbachia*-infected *Aedes aegypti* in Niterói, a city of half a million people in Rio de Janeiro state, where *Wolbachia* releases across 70% of the urban population in 2017-2019 were expanded to remaining populated areas in 2023. *w*Mel was durably established at ≥95% prevalence in *Ae. aegypti* populations throughout Niterói four years post-release, and up to seven years in the earliest release sites. This city-wide *Wolbachia* coverage provided sustained population-level protection against dengue throughout the five years post-intervention, including during the 2024 epidemic surge, averting an estimated three-quarters of the dengue case burden that would otherwise have been expected in Niterói in 2024.

## Introduction

Dengue has been a public health problem in the tropical world for decades. A global dengue surge in 2024 saw in excess of 14 million cases and 10 thousand deaths reported worldwide (1) – more than doubling the largest epidemics previously recorded in 2023 and 2019. The Americas region accounted for the vast majority of reported dengue cases and deaths in 2024 (1). Brazil was among the worst affected countries, reporting 6.6 million cases and 6200 deaths (2), and the projected economic impact on the country was estimated at US$3 billion well before the true magnitude of the epidemic was evident (3).

Arboviral disease prevention and control efforts in the Americas and elsewhere have long emphasised an intersectoral approach integrating epidemiological and entomological surveillance, chemical and environmental control of mosquito populations, together with effective clinical management (4). The success of these efforts in mitigating dengue epidemic spread is hampered by insecticide resistance, the limited feasibility of scaling and sustaining vector source reduction activities in complex urban environments, and limited funding for these recurrent activities within public health budgets. Vaccines for dengue and chikungunya have been authorised for use in several countries since 2022, and in February 2024 Brazil became the first country to launch a public dengue vaccination program, targeting 10 to 14 year-olds in the municipalities with highest case incidence (5). Brazil has also been at the forefront of evidence generation for *Wolbachia*-based arboviral disease control, a self-sustaining strategy that involves time-limited releases of *Aedes aegypti* mosquitoes carrying the endosymbiotic insect bacterium *Wolbachia*, which spreads into the local *Ae. aegypti* population and significantly reduces the mosquitoes’ capacity to transmit dengue and other viruses (6–10).

Following successful pilot releases of *w*Mel-strain *Wolbachia*-infected mosquitoes in the adjacent municipalities of Rio de Janeiro (11) and Niterói (12,13) in 2014 and 2015, respectively, phased expanded releases were conducted in the two cities between 2017 and 2019. Early monitoring showed evidence of significant reductions in dengue and chikungunya incidence in *Wolbachia*-treated areas in both cities, despite incomplete and spatially variable levels of introgression within the first 1-2 years post-release (13,14) – consistent with an accumulating body of evidence from randomised and non-randomised field trials in Australia, Asia and Latin America (15–20). Economic modelling studies in Brazil (21) and elsewhere (22,23) concur on the long-term cost-effectiveness of *Wolbachia-*based disease control under scenarios of scaled programmatic deployment in high-burden, populous cities, however challenges remain with financing the upfront costs of initiating and scaling *Wolbachia* release programs. A better understanding of the durability and long-term public health impact of *Wolbachia* deployments in different settings can help inform scale-up plans, including optimal integration with vaccination programs and integrated vector management efforts. The long-term durability of *w*Mel-*Wolbachia* in the field has been demonstrated a decade post-release in northern Queensland, Australia (24,25), and up to four and six years after the earliest releases in Colombia (26) and Indonesia (19), respectively. Here we report the long-term entomological and epidemiological outcomes of city-wide *Wolbachia* deployments in Niterói, a Brazilian city of half a million people, which demonstrate sustained public health benefits in the context of an unprecedented dengue outbreak in Brazil.

## Results

### City-wide coverage and long-term stability of Wolbachia in the Niterói Ae. aegypti mosquito population

Following the phased deployment of *w*Mel-infected *Ae. aegypti* throughout three-quarters of Niterói’s urban population between February 2017 and December 2019 (release zones 1 - 4; population 371,000) (13), releases of the same *w*Mel-infected *Ae. aegypti* line were expanded to the remaining urban areas of the city (zone 5; population 111,000) between November 2022 and July 2023 (**Figure 1A; Supplementary Table S1**). In zone 5, an estimated 21.3 million mosquitoes were released as adults from moving vehicles over 32 release weeks and the establishment of *w*Mel in the local *Ae. aegypti* population was monitored in larvae reared from ovitrap collections undertaken by municipal health authorities as part of routine vector surveillance activities (see Methods). *w*Mel prevalence in zone 5 was 51% (44 *w*Mel-positive of 86 larvae tested from 15 traps) by August 2023, 3 months post-release and 92% (22/24 larvae from 10 traps) at last monitoring in October 2023, 5 months post-release (**Figure 1B**).

**Figure 1:**
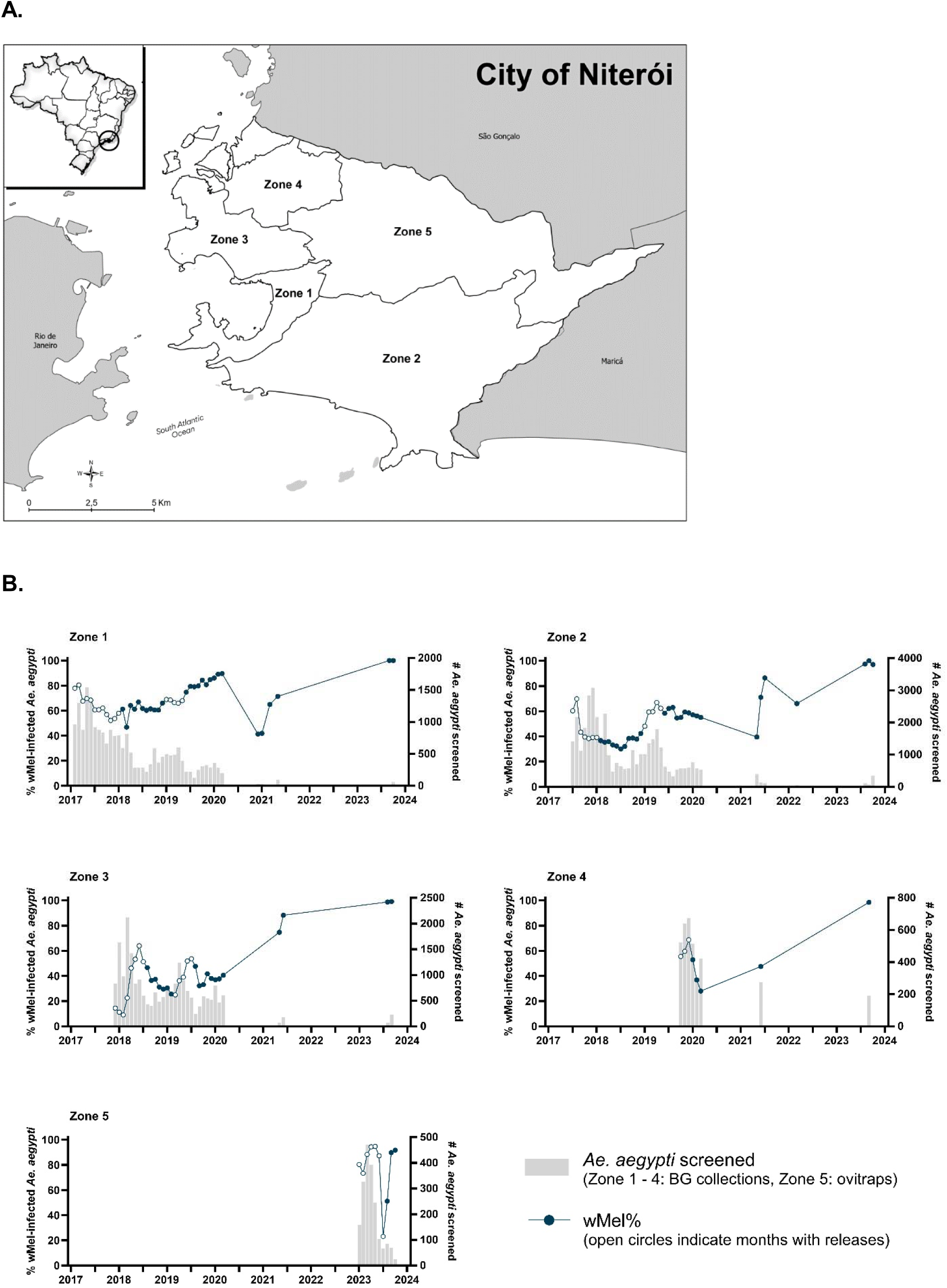
*w*Mel infection prevalence in *Aedes aegypti* mosquitoes collected from each release zone, during and after *Wolbachia* releases. A. Map of Niterói showing the release zones where *w*Mel-infected *Ae. aegypti* were released in February 2017 - January 2018 and January - May 2019 (Zone 1); June 2017 - January 2018 and January - May 2019 (Zone 2); November 2017 - July 2018 and March - July 2019 (Zone 3); September - December 2019 (Zone 4); and November 2022 - July 2023 (Zone 5). B. Circle markers represent the aggregate *w*Mel infection prevalence in each zone in each calendar month from January 2017 to October 2023. Open circles indicate months when *Wolbachia* releases took place in any part of that zone; filled circles are months with no releases. Bars show the number of *Ae. aegypti* tested for *w*Mel infection by qPCR. In zones 1 - 4 adult *Ae. aegypti* were collected with BG traps, and in zone 5 ovitraps were used to collect eggs that were reared to larvae for species identification and *w*Mel testing. Monitoring results with <20 *Ae. aegypti* tested were excluded from the graphs (Zone 1: n=4 observations; Zone 2: n=6; Zone 3: n=4; Zone 5: n=3).

Long-term monitoring of the invasion and stability of *w*Mel in the *Ae. aegypti* populations in the 2017-2019 release areas (zones 1-4) was conducted between 2021 and 2023, using BG Sentinel traps and mechanical aspirators to catch and screen adult mosquitoes from across the release areas, consistent with the initial monitoring up to March 2020 reported previously from these areas (13). At last monitoring in July - October 2023, four years after the end of releases, between 97% and 100% of the *Ae. aegypti* screened in each zone were *w*Mel-positive (**Figure 1B**. n=1096 total *Ae. aegypti* screened; 80 - 527 per zone).

This high level, durable wMel establishment was observed consistently throughout Niterói, with *w*Mel prevalence ≥95% in the *Ae. aegypti* population in every neighborhood in zones 1-4 that was monitored in 2023 (26 of 32 total neighborhoods), 44 - 53 months after the end of releases (**Figure 2**).

**Figure 2:**
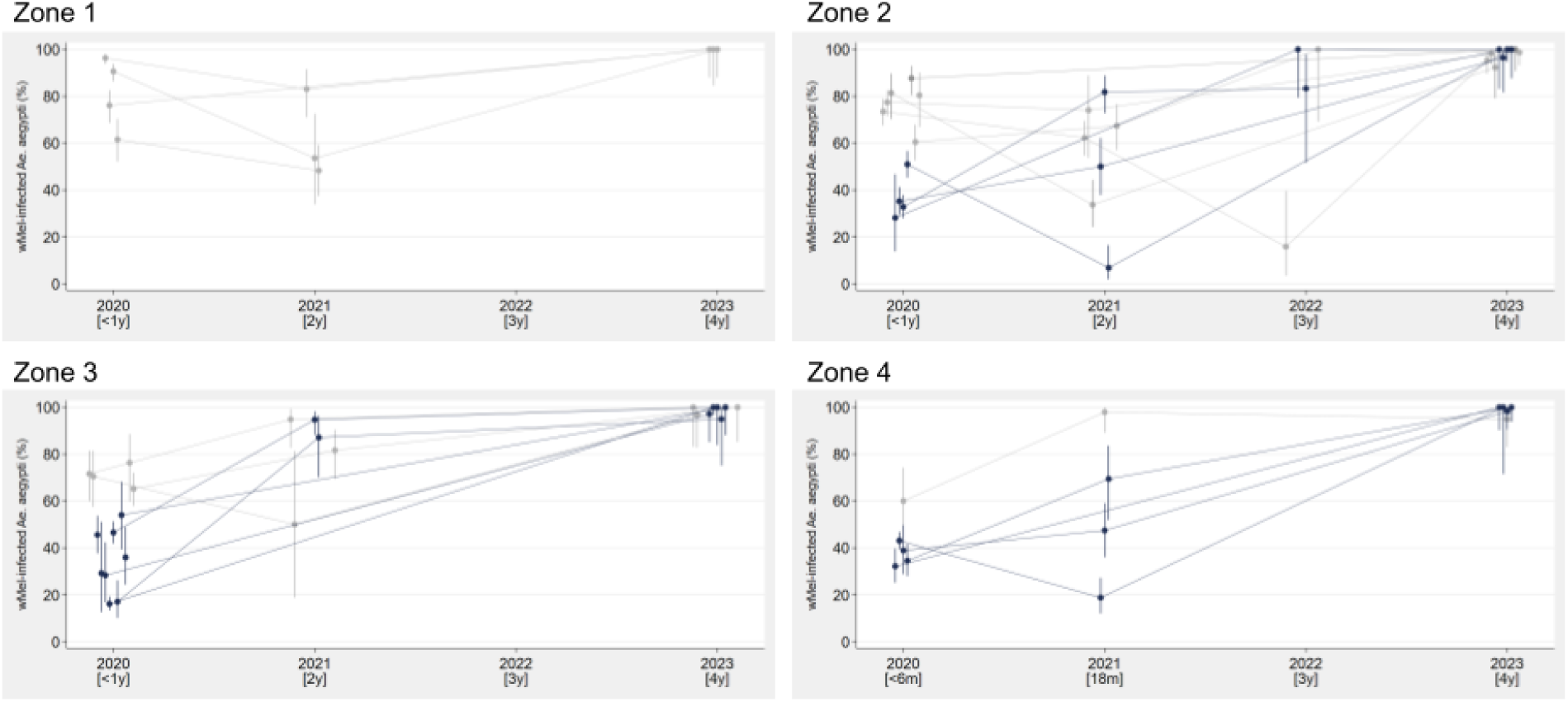
*Wolbachia* invasion profiles in individual neighborhoods during long-term monitoring up to four years post-release. *w*Mel-infected *Ae. aegypti* mosquito releases were completed in May (Zones 1 & 2), July (Zone 3) or December (Zone 4) 2019. Markers represent the percentage of adult *Ae. aegypti* collected by BG trap or aspiration that were positive for *w*Mel *Wolbachia* by PCR; vertical lines show the 95% confidence interval for the sample proportion, calculated by the Clopper-Pearson exact method. Collections were conducted across 1-3 months each year in each neighborhood, and aggregated by year to account for different collection months and relatively small sample sizes. Observations where <10 *Ae. aegypti* were screened from a neighborhood in a year were excluded. Monitoring in 2020 was between January and March, and these data have been reported previously (13). Of 32 neighborhoods in zones 1-4, all but two had *w*Mel% results in quarter 1 2020; 26 of these neighborhoods were monitored in 2023 (n>10 *Ae. aegypti*), and 18 have additional monitoring results from 2021 and/or 2022. Grey lines indicate neighborhoods where *w*Mel was already well-established (≥60%) in quarter 1 2020, within 6-12 months of releases, and black lines are neighborhoods where *w*Mel prevalence was <60% in short-term monitoring.

*Ae. albopictus* was detected throughout the city during the long-term monitoring period (**Supplementary Figure S1**), at a variable relative abundance compared to *Ae. aegypti*.

### Sustained suppression of dengue transmission in Niterói

Dengue notifications data was available for ten years (2007 - 2016) prior to the first *Wolbachia* mosquito releases in Niterói, during which time a total of 43,488 dengue cases were reported in the city: an average of 4349 notified cases per year [annual range 366 - 11,619], corresponding to 913 cases per 100,000 people per year [annual range 75 to 2396/100,000 people]. By comparison, in the five-year period October 2019 - September 2024 during which *Wolbachia* has been deployed throughout the entire urban area of Niterói, there were a total of 2193 dengue cases reported in the city: an average of 439 cases per year, corresponding to an average incidence of 91 cases per 100,000 people per year. Notably, 83% (n=1820) of those cases were reported in 2024 during a period of unprecedented high dengue case incidence in the Americas, including Brazil, following four years of near-zero dengue incidence in Niterói (**Figure 3**).

**Figure 3:**
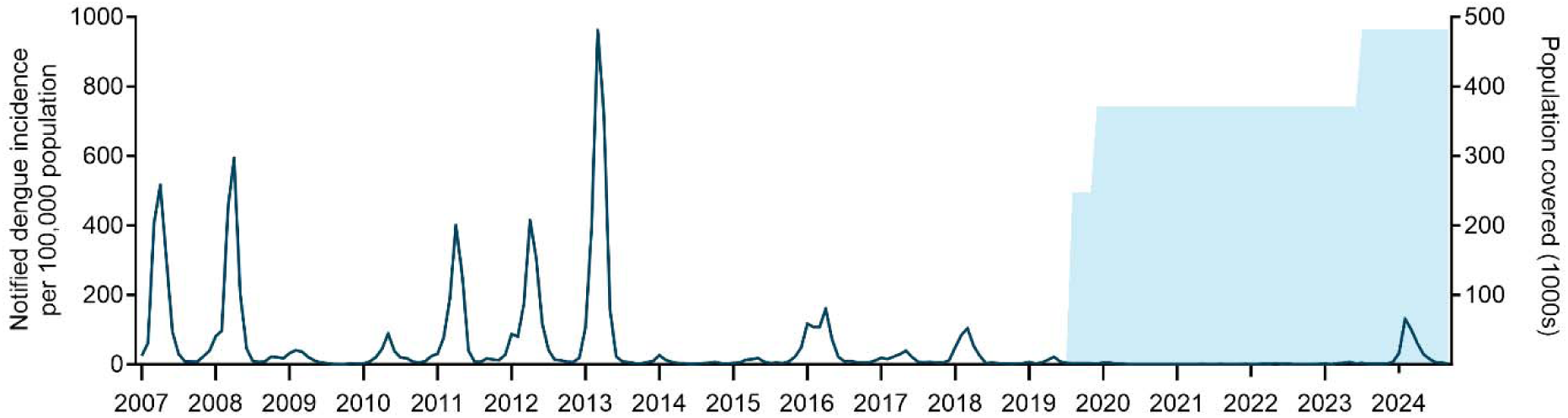
Dengue incidence in Niterói before, during and after phased *Wolbachia* mosquito releases. Dark blue line is the monthly incidence of dengue case notifications per 100,000 population in Niterói from January 2007 to September 2024. Blue shading indicates the cumulative residential population of the areas in which *Wolbachia* releases had been completed.

There was a near absence of chikungunya and Zika case notifications in Niterói over the five years post-intervention: a total of 131 chikungunya and 34 Zika cases were notified between October 2019 and September 2024 (average annual incidence 5.4 and 1.4 per 100,000 population, respectively) (**Supplementary Figure S2**).

Using interrupted time series (ITS) analysis to account for underlying temporal trends and the staggered implementation of the intervention across Niterói, notified dengue incidence was estimated to be 88.8% lower (95% confidence interval: 77.9 to 94.4%) following the completion of *Wolbachia* releases, compared to the 10-year pre-intervention period of 2007 - 2016. Dengue incidence was also significantly lower in the period during which releases were ongoing, compared to pre-release (90.2% reduction; 95%CI: 86.1 to 93.0%).

The inherent inter-annual and spatial variability in dengue epidemic cycles is a limitation in inferring intervention effects from a before-and-after analysis, or from comparisons with individual untreated municipalities. To inform an assessment of the dengue case incidence that could have been expected in Niterói in 2024 in the absence of the *Wolbachia* intervention - and thus the effectiveness of city-wide *Wolbachia* coverage in averting a larger outbreak in Niterói - we compared annual per capita dengue incidence in Niterói against the rest of Rio de Janeiro state and Brazil prior to, during and after *Wolbachia* deployments (2007 - 2024). Niterói has historically experienced high dengue incidence relative to other municipalities, ranking in the top 10 among the 28 large municipalities (≥100,000 population) in Rio de Janeiro state in all but one of the 12 years prior to the completion of area-wide releases in 2019. Dengue incidence in Niterói exceeded the Rio state average in ten of 12 years and exceeded the national average in seven of 12 pre-intervention years (**Figure 4; Supplementary Figure S3**). This ranking has changed dramatically following *Wolbachia* deployments, with annual dengue incidence in Niterói 44 - 89% lower than the state average and 88 - 98% lower than the national average in every year since 2020 and Niterói ranking among the lowest incidence cities in the state each year since 2022 **(Figure 4)**.

**Figure 4:**
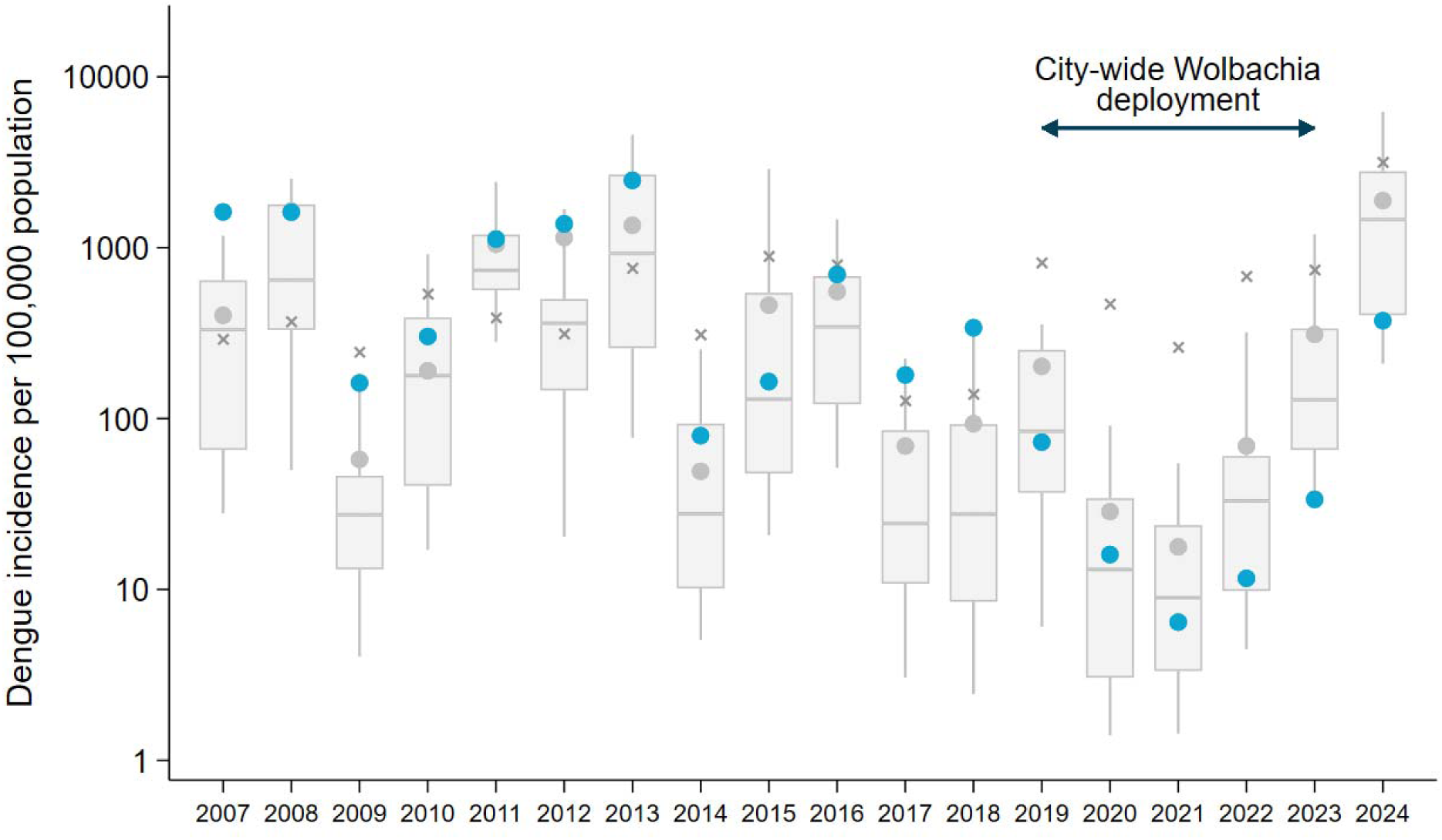
Annual dengue incidence in Niterói before, during and after *Wolbachia* deployment, compared to Rio de Janeiro state and nationally. The annual incidence of notified dengue cases per 100,000 population in Niterói (blue circles) is shown relative to the annual incidence among all cities (N=28) in Rio de Janeiro state with population >100,000. Box plots show the median and interquartile range of the incidence in the 28 cities, and spikes show the 10^th^ and 90^th^ percentiles. The aggregate annual incidence in all of Rio de Janeiro state (grey circles) and all of Brazil (crosses) is shown for each year. The timeline of phased city-wide roll-out of *Wolbachia* is indicated, including the release period (dotted line) and post-release period (solid line) in zones 1-4, covering 70% of the urban population, and the expansion of releases to the remaining urban areas (thicker line).

Using the median and 75th percentile of the 2024 dengue incidence among all the large cities in Rio de Janeiro state to project a range of dengue cases that could reasonably have been expected in Niterói in 2024 in the absence of *Wolbachia*, we estimate conservatively that the *Wolbachia* intervention averted at least 5242 to 11,660 dengue cases in Niterói in 2024, corresponding to a reduction in case burden of between 74% and 87%.

## Discussion

The *w*Mel strain of *Wolbachia* was durably established in local *Ae. aegypti* populations throughout the Brazilian city of Niterói by late 2023, making Niterói the first city in Brazil with citywide *Wolbachia* coverage. A four-year period of historic low dengue incidence in 2020 - 2023 followed *Wolbachia* deployment, before a relative increase in incidence was observed in Niterói in 2024 during an unprecedented dengue outbreak in Brazil and the region. However, case incidence in Niterói remained substantially lower than would be expected based on comparisons with historical dengue trends in Niterói and with all other large municipalities in Rio de Janeiro state and nationally. Our findings indicate that *Wolbachia* prevented at least three-quarters of the dengue case burden that may have otherwise occurred in Niterói in 2024, corresponding to thousands of cases averted. This extends existing evidence by showing that the public health benefits of successful area-wide establishment of *Wolbachia* are sustained even in a context of very high dengue transmission intensity.

A consistently high prevalence of *w*Mel (≥95%) was observed in the *Ae. aegypti* population in each of the 26 individual neighborhoods of Niterói monitored >4 years post-release, despite substantial initial variability of invasion levels within the first 12 months of post-release monitoring, and with no re-releases undertaken. In the 2023 expanded release area, material for monitoring *w*Mel invasion was collected using ovitraps instead of adult mosquito collections, in order to integrate field monitoring with the routine vector surveillance activities undertaken by municipal health department staff. A switch to ovitrapping was also implemented in 2024 for long-term monitoring of the earlier release areas, but the 2024 ovitrap data were excluded from our analyses due to very small sample sizes and limited spatial representativeness. While ovitrapping provides an opportunity to reduce costs by integrating *Wolbachia* monitoring with routine public health activities, additional work is required to validate the comparability of *w*Mel prevalence estimates from egg versus adult collections in the same location. Although a previous study in Brazil showed a high correlation between weekly *w*Mel *Wolbachia* frequency estimates based on screening of larvae obtained from eggs collected from ovitraps, and adult mosquito collections from BG-Sentinel traps (27); these were based on relatively large weekly sample sizes (approximately 200 individuals for each method per week) and high density of traps (125/km^2^ and 188/km^2^, BG-Sentinel and ovitraps, respectively). Large scale monitoring of *Wolbachia* establishment using ovitraps needs to take into consideration the minimum overall and maximum number of samples per trap for screening, the spatial distribution of egg collections, and the storage conditions for eggs and larvae prior to *Wolbachia* PCR screening.

The occurrence of reported dengue cases in an area where *Wolbachia* has been deployed is not unexpected for several reasons. Firstly, notified dengue cases in Brazil and most other endemic settings are probable cases, reported on the basis of a clinical case definition (28) and often without laboratory testing, meaning an unknown proportion are febrile illnesses of another aetiology. Secondly, cases are notified by place of residence, but may have acquired their infection in other locations. Thirdly, even where area-level monitoring shows very high *Wolbachia* prevalence, there may remain pockets of wild-type *Ae. aegypti* sufficient to sustain limited local dengue transmission, especially when overall mosquito abundance is exceptionally high due to climatic or other factors. Finally, the virus-blocking effect that *Wolbachia* (*w*Mel and *w*AlbB strain) confers on *Ae. aegypti* is not perfect and laboratory experiments have shown that mosquitoes can develop infectious saliva (‘breakthrough infection’) despite the presence of *Wolbachia*, especially when the viral load in the blood meal is very high, and with a higher probability for DENV-1 than for other serotypes (29,30). Nevertheless, mathematical modelling predicts a significant and sustained reduction in dengue incidence in human populations even with incomplete virus blocking or *Wolbachia* coverage in the mosquito population (30–32) reduction in the effective reproduction number during an outbreak (the average number of new infections arising from an infected individual). Consistent with this, our findings suggest that *Wolbachia* significantly reduced the peak of the epidemic curve and thereby the overall case burden in Niterói, but did not eliminate all transmission during a year of exceptionally high national transmission intensity. We note also the presence of *Ae. albopictus*, a competent vector of dengue viruses, throughout Niterói.

The challenge for assessing the real-world effectiveness of *Wolbachia* deployments from routine data sources across different settings lies in evaluating observed dengue case occurrence against what might have been expected in the absence of the intervention – i.e. the counterfactual – which is difficult to establish with certainty given the natural spatial and temporal variability in dengue epidemic cycles. These constraints are not specific to *Wolbachia*-based dengue control, they are common to many public health evaluations where a well-matched untreated control group is unavailable. In such scenarios, interrupted time series analysis - using multiple consecutive pre- and post-intervention observations in a single population and explicitly incorporating time - has been shown to be a powerful quasi-experimental study design for estimating intervention effect (33,34). Bernal et al (2019) (33) note the importance of acknowledging explicitly the limitations in controlling for confounding contemporaneous events, and in considering biological plausibility, magnitude of effect and consistency across settings in interpreting ITS results. The internal consistency in our estimates of *Wolbachia* intervention effect from uncontrolled ITS (within-population analysis) and from a comparison of annual dengue incidence in Niterói versus other Brazilian cities (between-population analysis) - as well as the external consistency with intervention effects reported in other settings and the biological plausibility given very high *w*Mel prevalence - provides reassurance that these findings are robust to potential confounding effects including secular trends, artefacts of healthcare-seeking or reporting behaviours during the Covid-19 pandemic, or other factors.

This reduction in notified case incidence attributable to *Wolbachia* deployments in Niterói translates into a considerable broader impact at individual, health system and societal levels. The annual direct and indirect costs of dengue in Brazil were estimated at between US$ 517 and 1688 million in the years 2009 – 2013 (35), and the average cost per dengue case has been estimated at US$ 532 in 2013 dollars (36). Hospitalisation status is not well-recorded in the publicly available SINAN dengue notifications data, but previous studies have reported hospitalisation rates of dengue cases in Brazil ranging from 3-4% up to 13-30% (35–37). Even at the lower end of this range, this indicates a substantial alleviation of dengue-related hospital admissions in Niterói in 2024 as well as the associated healthcare costs of both hospital and outpatient care, and the indirect household and societal costs of ill-health and care-giving.

In response to the rapid expansion in the population at risk of dengue in Brazil and in the frequency and magnitude of dengue epidemics, the Brazilian Ministry of Health published in January 2025 a revised national contingency plan for dengue, chikungunya and Zika which identifies the phased expansion of *Wolbachia* releases and other new vector control technologies as a priority activity, alongside the roll-out of dengue vaccination programs and the strengthening of capacity in surveillance, outbreak response and clinical case management (38). The scale of the dengue problem in Brazil and the region highlights the importance of an integrated approach that employs all available solutions. Monitoring of entomological and epidemiological outcomes of *Wolbachia* deployments in Brazil, including in cities where *Wolbachia* establishment has been less homogenous, are critical for a balanced evaluation of *Wolbachia* as a public health intervention. An ongoing randomised controlled trial of *Wolbachia* deployment in Belo Horizonte will add to the evidence-base (39). Similarly, evaluation of local dengue vaccination campaigns, and the comparative effectiveness and cost-effectiveness of *Wolbachia* and vaccination together or alone, will also be important in informing optimal strategies for integrating these new evidence-based interventions into disease control programs in Brazil and the many other countries facing a rising burden of dengue.

## Methods

### Ethics Statement

Approval to release *Wolbachia*-carrying *Ae. aegypti* mosquitoes into urban areas was obtained from three Brazilian governmental bodies: the National Agency of Sanitary Surveillance (ANVISA); the Ministry of Agriculture, Livestock and Supply (MAPA); and the Brazilian Institute of Environment and Renewable Natural Resources (IBAMA), which issued a Temporary Special Registry (Registro Especial Temporário (RET), nr. 0551716178/2017). Ethical approval was also obtained from the National Commission for Research Ethics (CONEP—CAAE 59175616.2.0000.0008).

### Mosquito Production

The *w*Mel-infected *Ae. aegypti* lines are maintained in controlled laboratory conditions. Immature stages for adult releases were reared at a density of approximately 2.75 larvae/ml and fed a diet of fish food: liver powder: yeast extract (4:3:1) (13). For zones 1-4, the production has been described previously (13). After adjustments and improvements, for Zone 5, when approximately 40% of larvae had pupated, the material were processed through a larvae/pupae separator and 220 pupae were placed in a release device, aiming to reach in the end 200 adult mosquitoes. These release devices were hexagon tubes approximately 60 mm in diameter and 170 mm in length, with a removable well on top. Adults were allowed to emerge for 5–6 days and were maintained on a 10% sugar solution for 12–36 hours prior to releases. The release tubes were then put into boxes and organized in vehicles for field releases.

### wMel deployment in Niterói

Releases of *w*Mel-*Ae. aegypti* throughout three-quarters of the urban population of Niterói (release zones 1-4) in 2017 - 2019 have been reported previously (13). For the expanded releases in the remaining urban areas of Niterói (zone 5), the density of release points was adjusted for the residential population in each neighborhood, with the aim of reaching a cumulative total of 200 mosquitoes per person. The release tubes containing an average of 200 mosquitoes, males and females, were distributed in routes and released from a vehicle, or on foot in areas where the vehicle couldn’t enter due to local restrictions. For these on-foot releases, a partnership with local vector control agents from the municipality was created and allowed the coverage of the whole territory. Weekly releases were conducted from November 2022 to July 2023, with between 28 and 32 release weeks per neighborhood.

### wMel monitoring

Long-term monitoring of *w*Mel prevalence in local *Ae. aegypti* populations in zones 1-4 used adult mosquitoes collected with BG Sentinel traps until 2020 as described previously (13), and then with aspirator collections from 2021 onwards (Improved Prokopack Aspirator Model 1419, John W. Hock Company, Gainesville, Florida, USA). In Zone 5, monitoring used mosquito eggs instead of adults, for the purpose of integrating field collections with the routine vector monitoring activities conducted by the Niterói municipal health authorities. *Ae. aegypti* eggs were sampled using an existing network of ovitraps (approximately one trap per 500 x 500 meter grid) containing a wood paddle where the females lay the eggs. Fortnightly, these traps were collected, the paddles were left to dry for 48 hours and then put into cups with water and fish food for hatching. After 4-5 days, L4 larvae were identified for species and maximum 20 larvae per trap were randomly selected for diagnostics, stored in 80% ethanol and then individually quantitative polymerase chain reaction (qPCR) processed for *w*Mel-strain *Wolbachia* detection (13).

### Dengue case notifications and population data

Dengue cases are notified to the Brazilian national disease surveillance system (SINAN) on the basis of a clinical case definition (28); only a minority of cases are laboratory confirmed. Line listed deidentified data was obtained from the SINAN system through the Health Secretariat of Niterói, on notified dengue cases resident in Niterói for the period January 2007 to September 2024 and on notified chikungunya and Zika cases for the period January 2015 to September 2024. Data was aggregated to monthly dengue case counts by *Wolbachia* release zone (1–5), based on cases’ neighbourhood of residence.

National data on annual notified dengue case numbers by municipality of residence for all of Brazil was also downloaded from the publicly accessible SINAN database (https://datasus.saude.gov.br/informacoes-de-saude-tabnet/) on 11 February 2025, for the years 2007 - 2024.

Data on the residential population of municipalities in Brazil was obtained from the 2000, 2010 and 2022 Brazilian censuses (IBGE: https://sidra.ibge.gov.br/pesquisa/censo-demografico). Census data from 2000 and 2010 was available by neighbourhood of residence for Niterói and was aggregated to estimate the population of each release zone, using 2000 data for the years 2007 - 2009 and 2010 data for the years 2010 - 2019. We estimated the release zone populations in 2020 - 2024 by applying the 2010 zone-level population distribution to the Niterói 2022 census population.

### Statistical methods

Per capita annual dengue incidence in Niterói was calculated as the annual number of notified cases per 1000 inhabitants. The *Wolbachia* intervention effect was estimated by interrupted time series analysis, implemented using mixed-effect negative binomial regression to model the monthly count of dengue case notifications in each release zone as a function of *Wolbachia* release status (before, during or after releases), with an offset for population size, calendar month as a fixed-effect covariate, and release zone modelled as a random effect. The model output is the dengue incidence rate ratio (IRR) in the post- or during-release periods compared with pre-release periods, adjusted for seasonality. Robust standard errors were used by specifying the vce(cluster *release_zone*) option in Stata to account for non-independence of observations within release zones.

## Data Availability

All data produced are available online at 10.26180/c.7756097

https://datasus.saude.gov.br/informacoes-de-saude-tabnet/

## Acknowledgments

The authors acknowledge the municipality of Niterói for their partnership and logistical support for this study and Fiocruz, for providing infrastructure for mosquito production and laboratory diagnostics. We are grateful to all WMP Brasil team for their dedication and to Eliane Moreira, Isabela Moreira and Mariana Magalhães for all project administration and logistical support.

