## Supplementary table and figures for "Long-term durability and public health impact of city-wide *w*Mel *Wolbachia* mosquito releases in Niterói, Brazil during a dengue epidemic surge"

**Supplementary Table S1. Summary of *w*Mel-*Ae. aegypti* releases and monitoring in Niterói.**

| **Zone** | **Population (2022)** | **Total Area (km^2^)** | **Release Area (km^2^)** | **# Release Periods** | **First Release** | **Last Release** | **Estimated mosquitoes released (millions)** | **# of Traps***  **(& after reduction)** | **Trap density* per km2 (& after reduction)** | **Start Monitoring** | **End Monitoring** |
| --- | --- | --- | --- | --- | --- | --- | --- | --- | --- | --- | --- |
| **1**^^^ | 23,592 | 8.5 | 3.5 | 2 | Feb 2017 | Dec 2019 | 2.64 | 138 (49) | 39 (14) | Feb 2017 | Oct 2023 |
| **2** | 68,246 | 51.3 | 18.9 | 2 | Jun 2017 | Jul 2019 | 12.84 | 302 (229) | 16 (12) | Jul 2017 | Oct 2023 |
| **3** | 177,722 | 12.2 | 9.4 | 3 | Nov 2017 | Jul 2019 | 12.61 | 169 | 18 | Dec 2017 | Aug 2023 |
| **4** | 101,119 | 10.9 | 8.1 | 1 | Sep 2019 | Dec 2019 | 6.17 | 140 | 17 | Oct 2019 | Sep 2023 |
| **5** | 111,070 | 50.5 | 16.2 | 1 | Nov 2022 | Jul 2023 | 21.29 | 105 | 6 | Jan 2023 | Sep 2024 |
| **Total** | 481,749 | 133.5 | 56.1 |  |  |  | 55.55 |  |  |  |  |

*In zones 1-4 *Wolbachia* monitoring used adult mosquitoes collected with BG traps (2017-2020) or aspirators (2021-2023), and in zone 5 used larvae reared from ovitrap collections.

^Release zone 1 includes the Jurujuba neighbourhood where pilot releases were conducted in 2015–16 (1), for all metrics except ‘Estimated mosquitoes released’ which includes only the expanded releases in zone 1 beginning in February 2017; the month that BG traps were first installed in zone 1 also excludes the pilot release period.

Note: release area comprises all urban or constructed areas in the zone, but excludes green non-constructed areas, which are less favourable habitats for *Ae*. *aegypti*. The number and density of BG traps was reduced in parts of zone 1 and zone 2 in order to reduce monitoring costs, once releases were completed and neighbourhood-level *w*Mel prevalence was *>*60% in 3 consecutive monitoring events measured at least 4 weeks after the conclusion of releases.

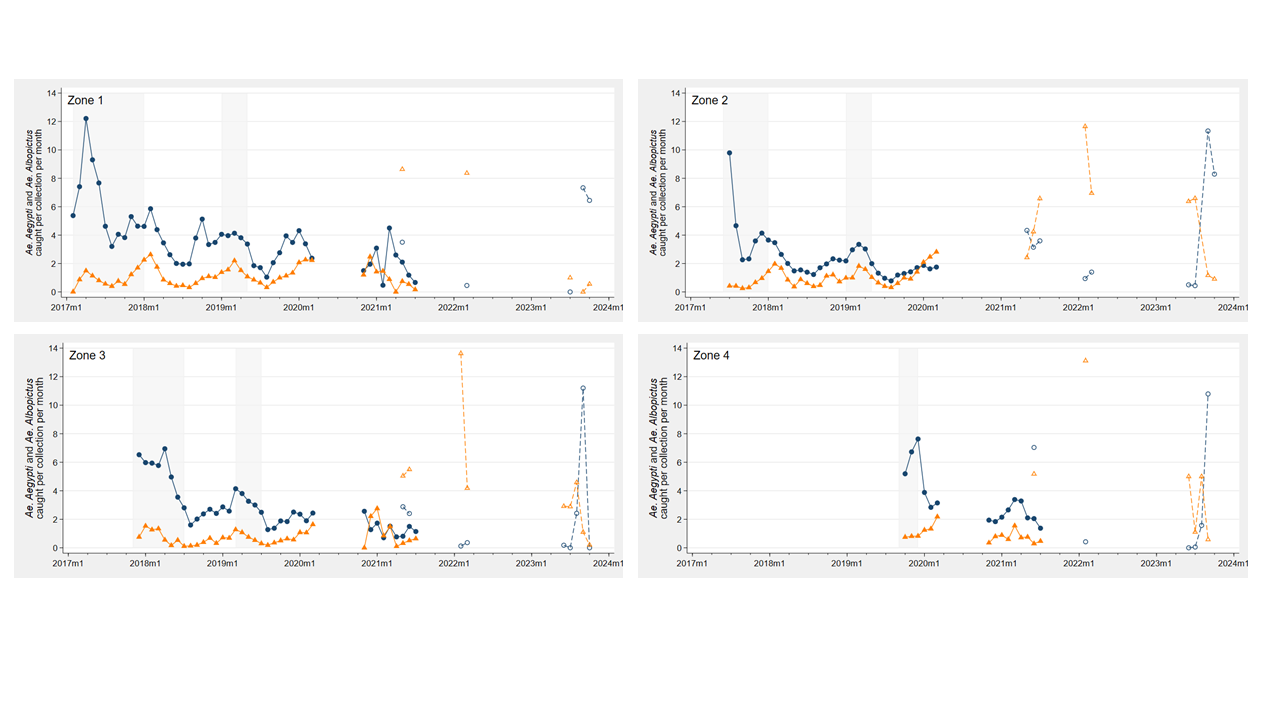

**Supplementary Figure S1: Relative abundance of *Ae. aegypti* and *Ae. albopictus* in long-term entomological monitoring in Niterói during and after *w*Mel-infected *Ae. aegypti* releases.** Counts of *Ae. aegypti* (blue lines with circle markers) and *Ae. albopictus* (orange lines with triangle markers) caught by BG traps (solid lines and filled markers) or by mechanical aspirators (dashed lines and open markers) were aggregated for each release zone each month. Release periods are indicated with grey shading. Zone 5 is excluded because all monitoring was done using ovitraps, not adult collections. One outlier observation is omitted from the zone 1 graph (23 *Ae. albopictus* in one aspirator collection in February 2022) and one from the zone 2 graph (583 *Ae. albopictus* in 30 aspirator collections [mean 19.53 per collection] in August 2023) to improve visibility of the remaining data.

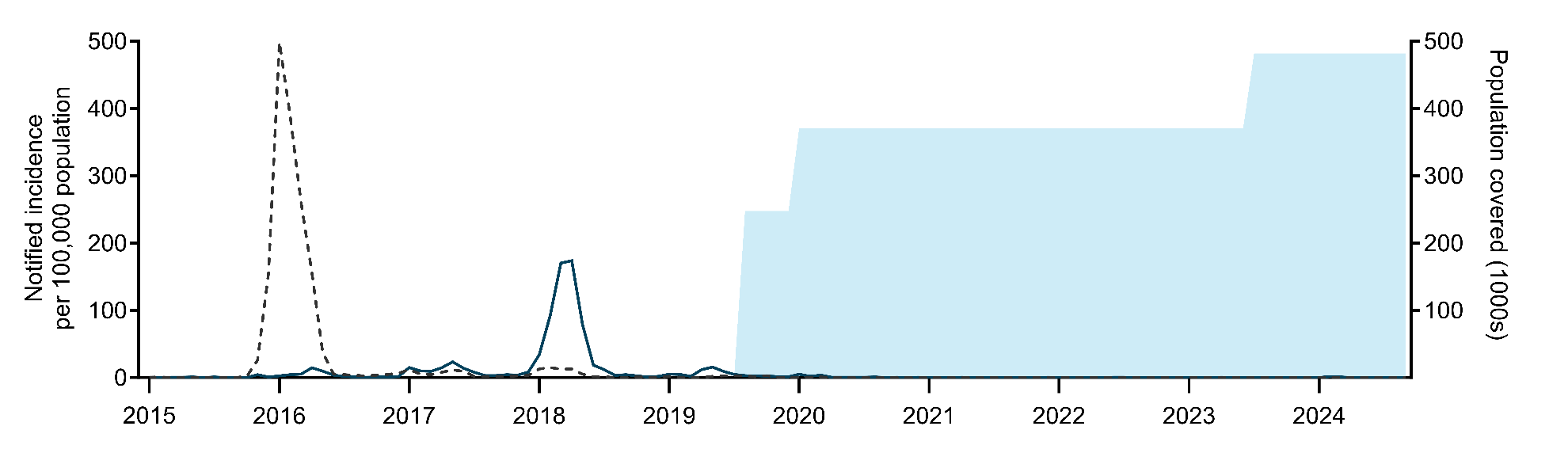

**Supplementary Figure S2**: Monthly incidence of notified chikungunya (solid line) and Zika (dashed line) cases per 100,000 population in Niterói, before, during and after phased *Wolbachia* mosquito releases (January 2015 to September 2024). Blue shading indicates the cumulative residential population of the areas in which *Wolbachia* releases had been completed.

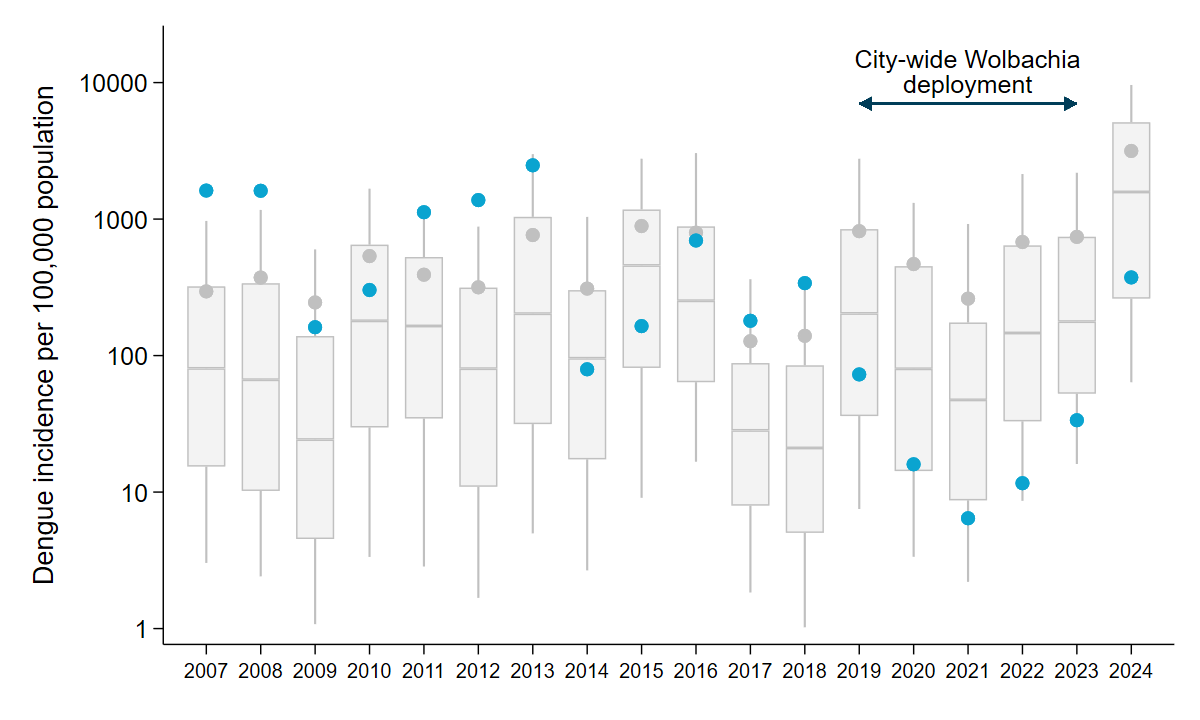

**Supplementary Figure S3: Annual dengue incidence in Niterói before, during and after *Wolbachia* deployment, relative to other dengue-affected cities in Brazil.** The annual incidence of notified dengue cases per 100,000 population in Niterói (blue circles) is shown relative to the annual incidence among all cities (N=316) in Brazil with population >100,000 and a non-zero number of dengue cases notified each year since 2007. Box plots show the median and interquartile range of the incidence in the 316 cities, spikes show the 10^th^ and 90^th^ percentiles of the incidence in the 316 cities, and grey circles show the aggregate incidence for all of Brazil. The timeline of phased city-wide roll-out of *Wolbachia* is indicated, including the release period (dotted line) and post-release period (solid line) in zones 1-4, covering 70% of the urban population, and the expansion of releases to the remaining urban areas (thicker line).
